# Selective interoception impairments and treatment effects in Borderline Personality Disorder

**DOI:** 10.1101/2025.11.04.25339393

**Authors:** Jella Voelter, Danilo Postin, Christina Müller, Mandy Roheger, André Schulz, Robin Bekrater-Bodmann, René Hurlemann, Dirk Scheele

## Abstract

**Background:** Patients with borderline personality disorder (BPD) suffer from severe emotional dysregulation and disturbances in body image and self-perception. Interoception, the processing and perception of internal body signals, is closely linked to emotional processing, but it remains unclear whether BPD impairs specific interoceptive facets and how these deficits respond to treatment.

**Methods:** We investigated the two key interoceptive facets, accuracy and attention, in 55 BPD patients and 31 healthy controls (HC) using self-report and objective measures before and after four-week residential Dialectical Behavior Therapy (DBT). Interoceptive accuracy and its metacognitive awareness were evaluated using a heartbeat discrimination task, while interoceptive attention was measured through questionnaires, intensity ratings, and both uni- and multivariate neural responses during a functional magnetic resonance imaging (fMRI) interoceptive attention task.

**Results:** Before DBT, BPD patients showed reduced self-reported interoceptive attention, which was associated with more interpersonal problems. Patients further exhibited higher similarity in activity patterns evoked by cardiac interoceptive and exteroceptive attention in the insular and dorsal anterior cingulate cortex. There were no significant group differences in behavioral or self-reported interoceptive accuracy or metacognitive awareness. However, behavioral interoceptive accuracy was impaired in BPD patients with more severe symptoms. After four-week residential DBT, self-reported interoceptive attention significantly improved, while other interoceptive facets showed no significant changes.

**Conclusions:** BPD involves disturbances in specific interoceptive facets that respond differently to treatment. Our findings support multifaceted assessments of interoception and the potential benefit of interoceptive attention training for all BPD patients, with additional accuracy training in more severe cases.

## Introduction

Borderline personality disorder (BPD) is characterized by severe emotional dysregulation, unstable interpersonal relationships, impulsivity, dissociations, and disturbances in body and self-image (1). Emotional processing has been closely linked to interoception, i.e., the perception and interpretation of inner body signals (2). Accordingly, interoception has gained increasing attention in contemporary psychiatric research, and deficits in interoception have been observed across various psychiatric disorders (3, 4). In contemporary models, interoception is conceptualized as a multifaceted construct (4). Murphy and colleagues (5) posit two central facets of interoception: interoceptive accuracy, the correspondence of perceived and actual body signals, and interoceptive attention, the degree to which interoception is the object of attention. Both can be assessed using objectifiable (behavioral or physiological) and self-report measures. While earlier studies typically focused on single interoceptive facets, recent studies highlight the importance of comprehensive, multidimensional assessment to reflect its complexity, identify possible disorder-specific interoceptive profiles, and improve treatment efficacy (5–7).

While current BPD treatment approaches already emphasize body awareness (8, 9), it remains unclear whether interoception contributes to BPD pathology or is effectively targeted by treatment, reflecting a limited understanding of its neural and behavioral mechanisms. An integrated biobehavioral model for BPD suggests that impaired interoception plays a central role in the complex psychosocial deficits in BPD by forming a triad with reduced emotional awareness and affective dysregulation that is influenced by genetic and environmental factors (10). However, few studies investigated BPD-associated impairments in specific interoceptive facets (11). For example, self-reported interoceptive attention was significantly decreased in patients with bulimia nervosa and comorbid BPD (12). Furthermore, heartbeat-evoked potentials (HEPs) as a physiological correlate of cardiac interoceptive attention were significantly reduced in BPD patients (13, 14). Reduced HEPs were associated with emotional dysregulation and structural alterations in the dorsal anterior cingulate cortex (dACC) and anterior insula (13), key regions for interoceptive processing (15). However, evidence on interoceptive accuracy in BPD is mixed: One study found that a patient group with various personality disorders, including BPD, exhibited reduced interoceptive accuracy in a heartbeat counting task (16). In contrast, another study reported no significant differences in interoceptive accuracy between BPD patients and HC in both heartbeat counting and heartbeat discrimination tasks (17). Interestingly, in a cross-sectional study, self-reported interoceptive attention mediated the association between childhood trauma and emotional dysregulation in BPD (18), suggesting that interoception may play a mechanistic role in BPD etiology. However, longitudinal studies are necessary to draw reliable conclusions about potential mechanisms of interoceptive dysfunctions in BPD.

Overall, the few existing studies on interoception in BPD converge mostly on the hypothesis that adverse and invalidating childhood environments may disrupt the integration of interoceptive and emotional signals, leading to deficits in interoceptive attention and an external attention bias, possibly contributing to the interpersonal and emotional dysfunction in BPD (11, 14, 18). Notably, reductions in cardiac interoceptive attention (i.e. HEPs) were not evident in remitted BPD patients (13), suggesting that intact interoception constitutes a resilience factor or that interoceptive impairment may be responsive to treatment. Dialectical Behavior Therapy (DBT) is currently the most empirically supported intervention for BPD, with meta-analytic evidence indicating moderate to large effect sizes in reducing self-injurious behaviors and improving psychosocial functioning (19). Originally derived from cognitive behavioral therapy (CBT), DBT incorporates a synthesis of acceptance- and change-oriented strategies, and targets core domains such as emotion dysregulation, interpersonal dysfunction, behavioral impulsivity, and mindfulness. The mindfulness component of DBT facilitates nonjudgmental awareness of internal experiences, which may contribute to enhanced interoceptive abilities, particularly when integrated with body-oriented therapeutic practices (8, 9, 20). Neuroimaging and psychophysiological studies have demonstrated that mindfulness practices can modulate neural substrates underlying interoception, including activity in the insula and anterior cingulate cortex (21–23). Furthermore, emerging evidence suggests small to moderate improvements in interoceptive attention and trauma-related symptomatology following mindfulness-based interventions (24).

In summary, evidence of impaired interoception in BPD remains scarce and multifaceted assessments and the investigation of treatment effects are missing. In the present study, we address these gaps by assessing interoceptive accuracy and attention with objectifiable measures (accuracy: heartbeat discrimination; attention: neural responses to interoceptive attention using functional magnetic resonance imaging; fMRI)), and self-reports (accuracy: confidence ratings; attention: intensity ratings and questionnaire scores (Multidimensional Assessment of Interoceptive Awareness, Version 2; MAIA-2)), in BPD patients before and after four weeks of a residential DBT program. Our pre-registered hypotheses stated that BPD patients, compared to HC, would exhibit impaired interoceptive abilities across all facets and altered activation in key interoceptive brain regions: the insular cortex, amygdala, dACC, and ventromedial prefrontal cortex (vmPFC) during interoceptive attention at the pre-measurement (i.e. before the DBT). Moreover, we explored associations with interpersonal problems, symptom severity, and childhood maltreatment. Finally, we hypothesized that BPD-associated impairments would be reduced after DBT treatment and that these treatment effects would be more pronounced in patients with stronger symptom reduction.

## Methods and Materials

### Study design and participants

The study design was registered at ClinicalTrials.gov (NCT04770038), and the analysis plan was pre-registered before conducting any analyses (https://osf.io/htf2n). A total of 69 BPD patients on a waiting list for DBT were recruited at the Karl-Jaspers-Klinik in Bad Zwischenahn, Germany. Trained clinical psychologists at the DBT outpatient clinic confirmed the BPD diagnoses through interviews as part of their clinical work. Prior to DBT, data from 55 BPD patients were compared to 31 HC without any psychiatric illness (see **Supplementary Information (SI)** for exclusion criteria, comorbidities, and medication of the patients). The analysis of DBT effects included 37 BPD patients and 31 HC, as nine BPD patients did not receive residential treatment during the study period. Since residential DBT is not an acute intervention, patients often wait several months before admission. Once admitted to DBT, two BPD patients (∼3.6%) dropped out of the study, and seven (∼12.7%) discontinued DBT before the second measurement (**Supplementary Fig. 1**). The sample size was based on an a-priori power analysis (**SI Power Analysis**). We aimed for an allocation ratio of 2:1 (patients:controls) based on the assumption of a 50% treatment response rate in BPD patients. Furthermore, we assumed a higher drop-out rate in BPD patients (20-25%) than in HC (10-15%), allowing us to investigate treatment effects in the longitudinal comparisons with sufficient statistical power. For demographic and clinical characteristics, see **Supplementary Tab. 1-3**. The study was approved by the University of Oldenburg medical ethics committee (2020-101) and conducted in accordance with the Declaration of Helsinki. All participants provided written informed consent.

### Residential DBT program

The residential DBT program at the Karl-Jaspers-Klinik is based on the inpatient DBT, established in 1995, and proven to be effective (9). The inpatient DBT program itself was adapted from the original outpatient DBT developed by Marsha Linehan (8). BPD patients register themselves at the DBT outpatient clinic and are invited to a first interview where a detailed assessment and discussion of life circumstances and treatment options takes place. This is followed by a pre-inpatient group, where treatment goals are discussed. The residential DBT program at the Karl-Jaspers-Klinik is structured into three modules, each lasting four weeks, with a varying outpatient practice phase in between. During these phases, patients are advised to apply the skills they have learned during therapy to their everyday lives. The first DBT module at the Karl-Jaspers-Klinik prioritizes the reduction of suicidal and parasuicidal behaviors, enhancing stress tolerance, managing cravings, and addressing dissociations. This module can function as a standalone treatment unit. Following this, the second module focuses on understanding and regulating emotions. The last module provides the opportunity to enhance interpersonal skills, foster self-esteem, and enhance overall quality of life. The treatment sessions include both group and individual therapy. During the group sessions, patients from all three modules participate together. The group sessions focus on psychoeducation, mindfulness and skills training, as outlined in the DBT skills training manual (25). The skills training forms the core of the DBT program. Patients learn strategies to effectively distract and calm themselves during high-stress situations, how to address interpersonal problems, and how to better perceive and regulate emotions through mindfulness. On the weekend, patients typically return home to engage in a stress trial in their usual environment. Please refer to the **SI Residential DBT program** for a more detailed description of the treatment components. To investigate DBT effects, BPD patients were measured before (pre-measurement) and after one module (i.e., four weeks) of the residential DBT program (post-measurement). The HC underwent the same measurements but with no intervention between pre- and post-measurement.

### Psychological and clinical assessments

The Borderline Symptom List-23 (BSL-23) (26) was utilized to evaluate the severity of BPD symptoms, the Childhood Trauma Questionnaire (CTQ) (27) was used to assess childhood trauma, and the Inventory of Interpersonal Problems (IIP) was administered to measure severity of interpersonal problems (28). BPD patients were subdivided into treatment responders and non-responders based on the Reliable Change Index (RC) (29) (**SI Psychological and Clinical Assessments**).

### Interoceptive accuracy

Interoceptive accuracy was measured by an adapted version of a heartbeat discrimination task (30) (**Fig. 1A**). Participants’ heartbeats were monitored using an electrocardiogram (ECG) (BIOPAC Systems, Inc.). Simultaneously, a sequence of five tones was presented, and participants had to indicate whether the tones were synchronous (R-wave + 250±75ms) or asynchronous (R-wave + 750±75ms) with their heartbeat. After each trial, participants rated their confidence in their judgment on a visual analogue scale (VAS) ranging from 0 (not confident at all) to 10 (very confident). Behavioral interoceptive accuracy was assessed based on performance, whereas confidence ratings were used as self-reported indicators. Furthermore, metacognitive sensitivity (i.e., how well a person can recognize the accuracy or inaccuracy of their judgments), and efficiency (i.e., metacognitive sensitivity relative to task performance) were calculated to evaluate metacognitive interoceptive awareness (31).

**Fig 1.**
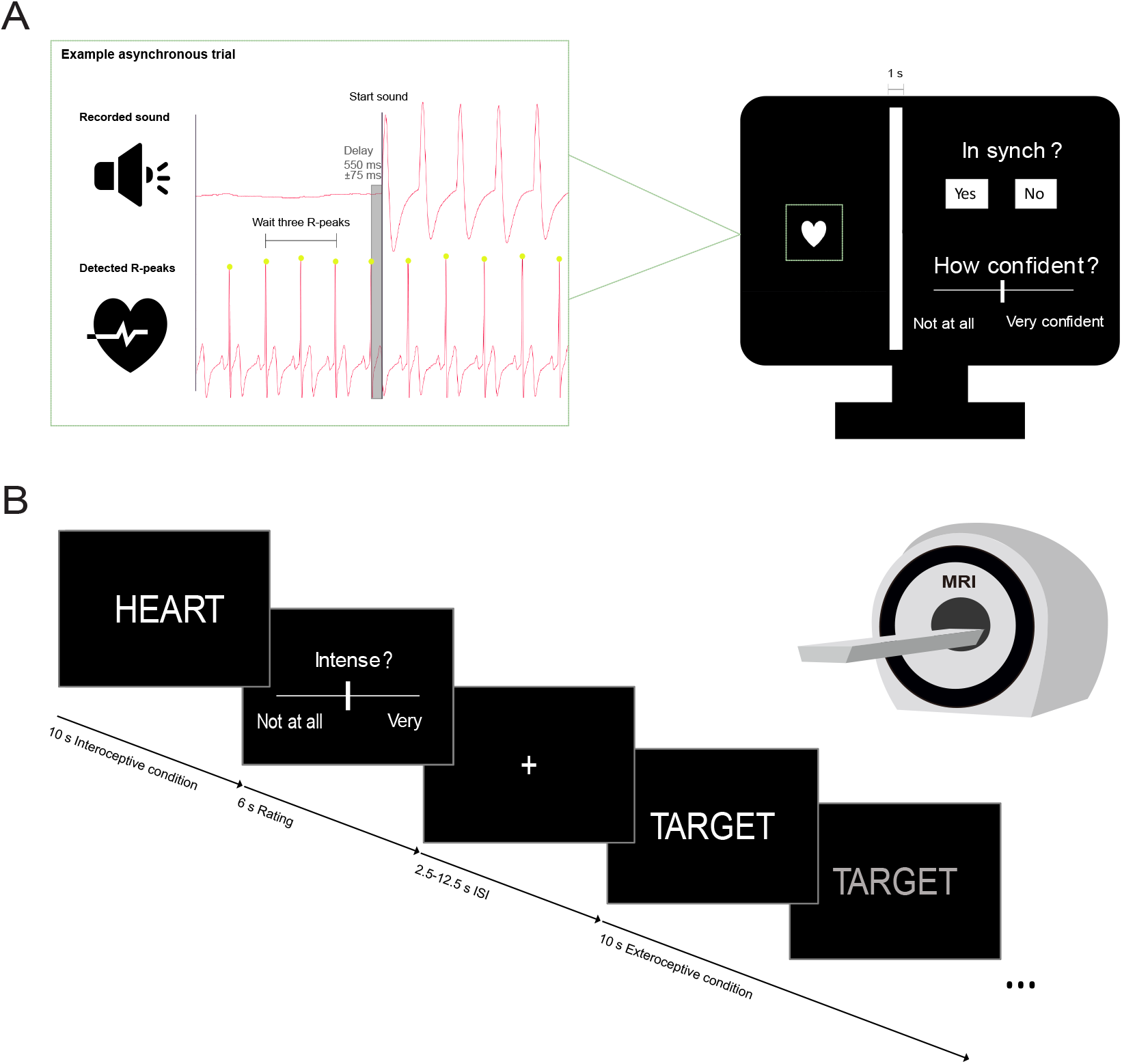
Heartbeat discrimination task **(A)**. Participants listened to a sequence of five tones while their heartbeat was monitored via electrocardiogram (ECG). Each tone sequence was triggered after detecting three consecutive, artifact-free heartbeats. During the tone sequence, a heart symbol was presented on the screen, and tones were either presented synchronous (250 ± 75 ms delay) or asynchronous (550 ± 75 ms delay) with the individual heartbeats. After each sequence, participants decided whether the tones were synchronous or asynchronous with their heartbeat and rated their confidence in the decision using a visual analog scale (VAS). The task consisted of 40 synchronous and 40 asynchronous trials. Task design of the visceral interoceptive awareness task **(B)**. During interoceptive conditions, the words “HEART” or “STOMACH” appeared on the screen, directing participants to focus on their heart or stomach, respectively. In the exteroceptive condition (“TARGET”), participants focused on the gradual shift in the word’s color from white to gray. In half of the trials, participants rated the perceived intensity of the trial on a VAS.

### Interoceptive attention

Self-reported interoceptive attention was assessed using the MAIA-2 (32). As a physiological indicator of interoceptive attention participants underwent an adapted version of an fMRI visceral interoceptive awareness (VIA) paradigm (33) (**Fig. 1B**). In interoceptive conditions, the word “HEART” or “STOMACH” was presented on the screen, cueing participants to focus on their heart or stomach, respectively. In the exteroceptive condition (“TARGET”), participants attended to gradual color changes of the presented word from white to gray. In half of the trials, participants rated the intensity of perceptions from their heart, stomach, or the color change of the word target on a VAS from 0 (not at all intense) to 10 (very intense).

### MRI data acquisition and preprocessing

MRI data were acquired using a 3T Siemens Prisma MRI scanner (Siemens AG, Erlangen, Germany) with a 64-channel head coil. High-resolution anatomical images were measured with a T1-weighted 3D MP-RAGE sequence. A T2*-weighted echoplanar multiband sequence with a multiband acceleration factor of 4 (34) was used to measure neural responses during interoceptive attention. fMRI data were preprocessed using the standardized pipeline *fMRIPrep* 20.2.1 (35).

### Behavioral data analysis

Behavioral interoceptive accuracy, metacognitive sensitivity, and efficiency were examined by calculating d’, meta-d’, and the meta-d’/d’ ratio (36), respectively. Analyses of Covariance (ANCOVAs) were used to compare questionnaire and behavioral data between groups, while repeated-measures ANCOVAs were performed to examine treatment effects within BPD patients. Moderation analyses were conducted to determine whether childhood trauma, BPD symptom severity, changes in BPD symptom severity, or treatment response moderated group differences (BPD vs. HC) or treatment effects (Pre vs. Post) in self-reported interoceptive attention, behavioral and self-reported interoceptive accuracy, and metacognitive efficacy. Furthermore, we employed correlational analyses using Pearson’s correlations to examine the relationships between these measures and interpersonal problems (IIP scores). Mixed-design ANCOVAs were conducted to compare changes in interoceptive indices in BPD patients to changes in the HC group. For all interoception measures, sex, age, Body Mass Index (BMI), and psychotropic medication use were included as covariates in the analyses. DBT module was included as a covariate for analyses investigating within-subject effects in BPD patients. Statistical significance was assessed at *p*<0.05, and post-hoc comparisons were corrected for multiple comparisons using Bonferroni-Holm correction (*p*_cor_).

### fMRI data analysis Univariate approach

The fMRI analysis was conducted in SPM12 and included sex, age, BMI, hunger, the urge to urinate, psychotropic medication use, and prescan inner tension as covariates. DBT module was included as a covariate for analyses investigating within-subject effects in BPD patients. All analyses were carried out using a single Region of Interest (ROI) mask comprising the insular cortex, amygdala, dACC, and vmPFC. To test whether group effects (BPD vs. HC) on neural responses before treatment were moderated by childhood trauma or BPD symptom severity, t-tests were calculated with an additional interaction term (e.g., group × CTQ scores). Likewise, we tested whether changes in symptom severity and treatment response moderated differences between the pre- and post-measurement. Significance was assessed at peak level with *p*<0.05, family-wise error (FWE) corrected.

### Multivariate approach

Representational Similarity Analyses (RSAs) were conducted on the individual ROIs, using the beta maps from the first-level analyses. The extracted Representational Similarity Matrices (RSMs) displaying Fisher z-transformed Pearson’s correlation coefficients between the individual conditions (heart, stomach, target) were compared between groups using bootstrapped two-sample t-tests and within-groups using bootstrapped paired-tests. Significance was assessed with *p*<0.05, Bonferroni-Holm corrected to adjust for the number of ROIs (*p*_cor_).

For fMRI and behavioral analyses details, see **Supplementary Methods**.

## Results

### Group differences prior to DBT

#### Symptom severity, childhood maltreatment and interpersonal problems

Before the treatment, BPD patients exhibited a high symptom load (BSL-23 score M±SD: 1.98±0.87) (37) that was significantly different from the ratings of HC (0.20±0.16; main effect of group: *F*_(1,81)_ =139.93, *p*<0.0001, η_p_^2^=0.63). Likewise, BPD patients reported significantly more severe childhood maltreatment (CTQ score: 65.00±21.00) and a higher degree of interpersonal problems (IIP score: 2.16±0.47) than HC (CTQ score: 35.06±10.03; *F*_(1,81)_ =64.13, *p*<0.0001, η_p_^2^=0.44; IIP score: 0.98±0.40; *F*_(1,81)_ =138.32, *p*<0.0001, η_p_^2^=0.63).

#### Interoceptive accuracy

BPD patients and HC did not differ significantly in their behavioral and self-reported interoceptive accuracy (**Fig. 2A-B**), metacognitive sensitivity, or efficiency (all *p*-values>0.05). However, a significant moderation effect of group × BSL-23 scores (*F*_(1,58)_ =9.03, *p*=0.004, η_p_^2^=0.13) on behavioral interoceptive accuracy shows that group differences in interoceptive accuracy become evident with increasing symptom severity. Johnson-Neyman intervals indicate that BPD (compared to HC) has a significant negative effect on behavioral interoceptive accuracy for a mild symptom severity of ≥0.47 (**Fig. 2C**).

**Fig 2.**
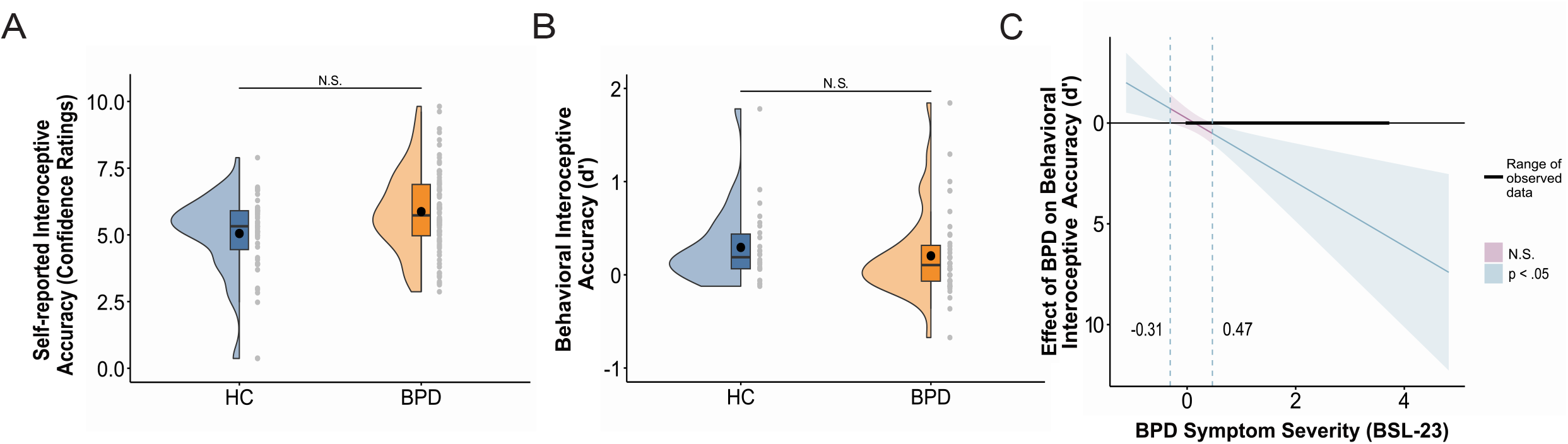
Self-reported and behavioral interoceptive accuracy at baseline. BPD patients (n=40) did not differ significantly from HC (n=26) in their self-reported (confidence ratings, **A**) or behavioral (d’, **B**) interoceptive accuracy. The Johnson-Neyman plot evaluates how symptom severity moderates the group difference (BPD vs. HC) in behavioral interoceptive accuracy. The interval in which the group difference is statistically significant is colored blue. Lower (−0.31) and upper (0.47) Johnson-Neyman intervals (dotted lines) reveal that BPD (compared to HC) has a significant negative effect on behavioral interoceptive accuracy from a symptom severity of 0.47 and above (**C**). Range of observed data for BPD symptom severity were BSL-23 scores of 0 (HC) to 3.70. In the boxplot, the line dividing the box and the black dot represent the median and mean of the data. The ends represent the upper/lower quartiles and the extreme lines represent the highest and lowest values excluding outliers. Abbreviations: BPD, borderline personality disorder; HC, healthy controls; N.S., not significant.

#### Interoceptive attention

BPD patients exhibited lower self-reported interoceptive attention (MAIA-2 score: 2.10±0.53; **Fig. 3A**) than HC (MAIA-2 score: 2.87±0.66; *F*_(1,78)_=34.52, *p*<0.0001, η_p_^2^=0.31), which correlated negatively with interpersonal problems in BPD patients (*r*_(51)_=-0.47, *p*_(cor)_=0.0007) and across both groups (*r*_(82)_=-0.62, *p*_(cor)_<0.0001), but not in HC (*r*_(29)_=-0.25, *p*_(cor)_=0.18; **Fig. 3B**). In the VIA task, interoception compared to the exteroceptive control condition resulted in significant activation within the bilateral insular cortex (right: peak MNI coordinates (x,y,z): 36, 8, 12; *t*_(24)_=5.71; *p*_FWE_ = 0.023, left: MNI: −34, 6, 4; *t*_(24)_=6.73; *p*_FWE_ =0.002; MNI: −38, −20, 0; *t*_(24)_=5.69; *p*_FWE_=0.025) and the right dACC (MNI: 2, 4, 48; *t*_(24)_=5.76; *p*_FWE_ =0.021) in HC. However, the univariate approach revealed no significant group differences during physiological interoceptive attention for any contrast (all *p*_FWE_**-**values>0.05; **Fig. 4A**) and intensity ratings did not differ significantly between BPD patients and HC (all *p*-values>0.05; **Fig. 4B**). Interestingly, the multivariate approach identified significant group differences in neural representations of interoceptive attention compared to exteroceptive attention in the insular cortex (**Fig. 4C**) and the dACC (**Fig. 4D**). Specifically, BPD patients exhibited greater similarity in neural activity patterns between interoceptive attention to the heart and the exteroceptive condition, characterized by significantly reduced negative correlations compared to HC in both regions (insular cortex: BPD: *r*_(48)_=-0.51, HC: *r*_(29)_=-0.61, *t*_(79)_=3.18, *p*_(cor)_=0.008, *d*=0.69; dACC: BPD: *r*_(48)_=-0.49, HC: *r*_(29)_=-0.61, *t*_(79)_=2.46, *p*_(cor)_=0.048, *d*=0.56). There were no significant group differences in neural representations of interoceptive attention to the stomach compared to the exteroceptive condition in the insular cortex and the dACC (all *p*_(cor)_-values>0.05).

**Fig 3.**
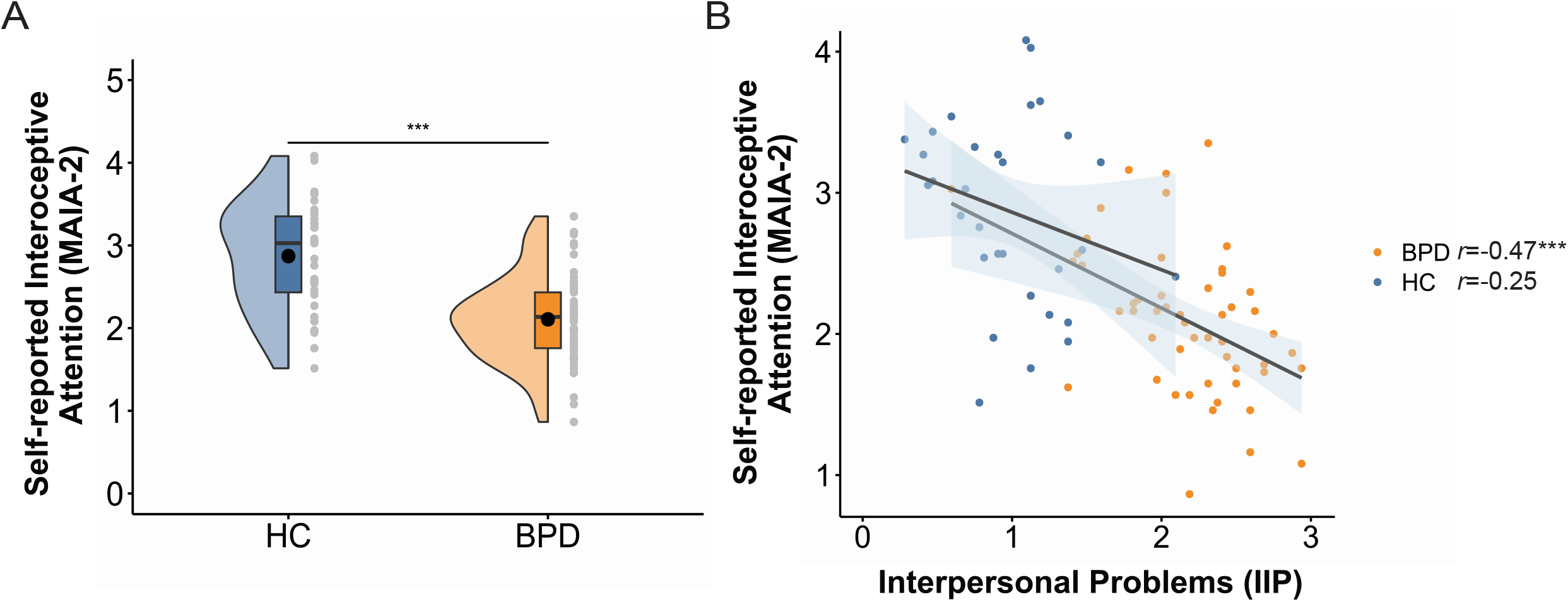
Self-reported interoceptive attention and interpersonal problems at baseline. Patients with BPD (n=53) compared to HC (n=31) showed significantly lower self-reported interoceptive attention (MAIA-2 scores, **A**). Self-reported interoceptive attention correlated negatively with severity of interpersonal problems (IIP scores) in BPD patients and across both groups (**B**). In the boxplot, the line dividing the box and the black dot represent the median and mean of the data. The ends represent the upper/lower quartiles and the extreme lines represent the highest and lowest values excluding outliers. The black lines in panel B represent the regression lines, and the shaded areas indicate the 95% confidence intervals. Abbreviations: BPD, borderline personality disorder; HC, healthy controls; \*\*\**p*<0.001. A Bonferroni-Holm correction was applied to adjust for multiple comparisons.

**Fig. 4:**
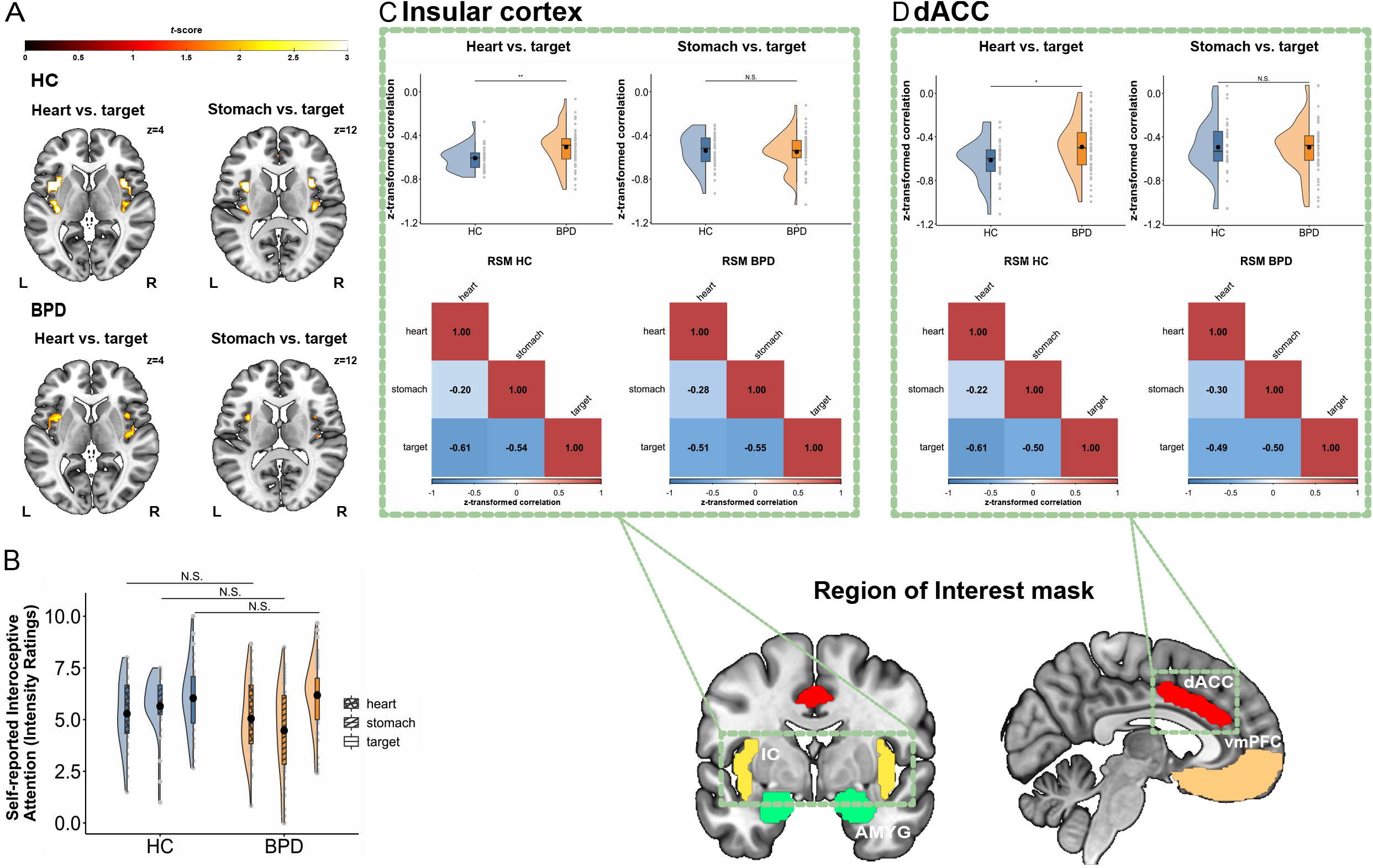
Intensity ratings of and neural responses to interoceptive attention at baseline. Patients with BPD (n=50) compared to HC (n=31) did not differ significantly in neural responses to interoceptive attention in the univariate analysis (**A**) nor in the intensity ratings (**B**). Representational Similarity Analyses results are shown as correlation matrices of multivariate neural patterns evoked by interoceptive (heart, stomach) and exteroceptive (target) attention in the insular cortex (**C**) and the dACC (**D**) for both BPD patients and HC. In both regions, BPD patients exhibited higher similarity in neural activity patterns between interoceptive attention to the heart and the exteroceptive condition compared to HC. No significant group differences were observed when comparing interoceptive attention to the stomach with exteroceptive attention in either region. For illustration purposes, the threshold for the univariate analysis was set to *p*<0.05 uncorrected. In the boxplot, the line dividing the box and the black dot represent the median and mean of the data. The ends represent the upper/lower quartiles and the extreme lines represent the highest and lowest values excluding outliers. Abbreviations: BPD, borderline personality disorder; HC, healthy controls; RSM, representational similarity matrix; IC, insular cortex; AMYG, amygdala; dACC, dorsal anterior cingulate cortex; vmPFC, ventromedial prefrontal cortex; \*\**p*<0.01.; \**p*<0.05; N.S., not significant. A Bonferroni-Holm correction was applied to adjust for multiple comparisons.

#### Moderation effects

There were no further significant moderation effects of symptom severity (BSL-23 scores) or childhood trauma (CTQ scores) on baseline group differences. Likewise, correlations with interpersonal problems (IIP scores) revealed no further significant associations.

### Investigation of DBT effects

#### Symptom severity, treatment response and interpersonal problems

After four weeks of a residential DBT program, BPD symptom severity showed significant improvement (main effect of time: *F*_(1,36)_ =37.75, *p*<0.0001, η_G_^2^=0.24) with a decrease from a high (2.10±0.82) to a moderate symptom load (1.22±0.83). Out of 37 patients with longitudinal data, 15 (41%) were classified as responders and 22 (59%) as non-responders. Furthermore, interpersonal problems significantly decreased after treatment in BPD patients (pre: 2.24 ± 0.17, post: 2.03 ± 0.27; *F*_(1,36)_ =8.03, *p*<0.007, η_G_^2^=0.05).

#### Interoceptive accuracy

Self-reported and behavioral interoceptive accuracy (**Fig. 5A-B**), metacognitive sensitivity, and efficiency did not improve significantly after treatment within BPD patients and compared to changes in HC (all *p*-values>0.05).

**Fig. 5.**
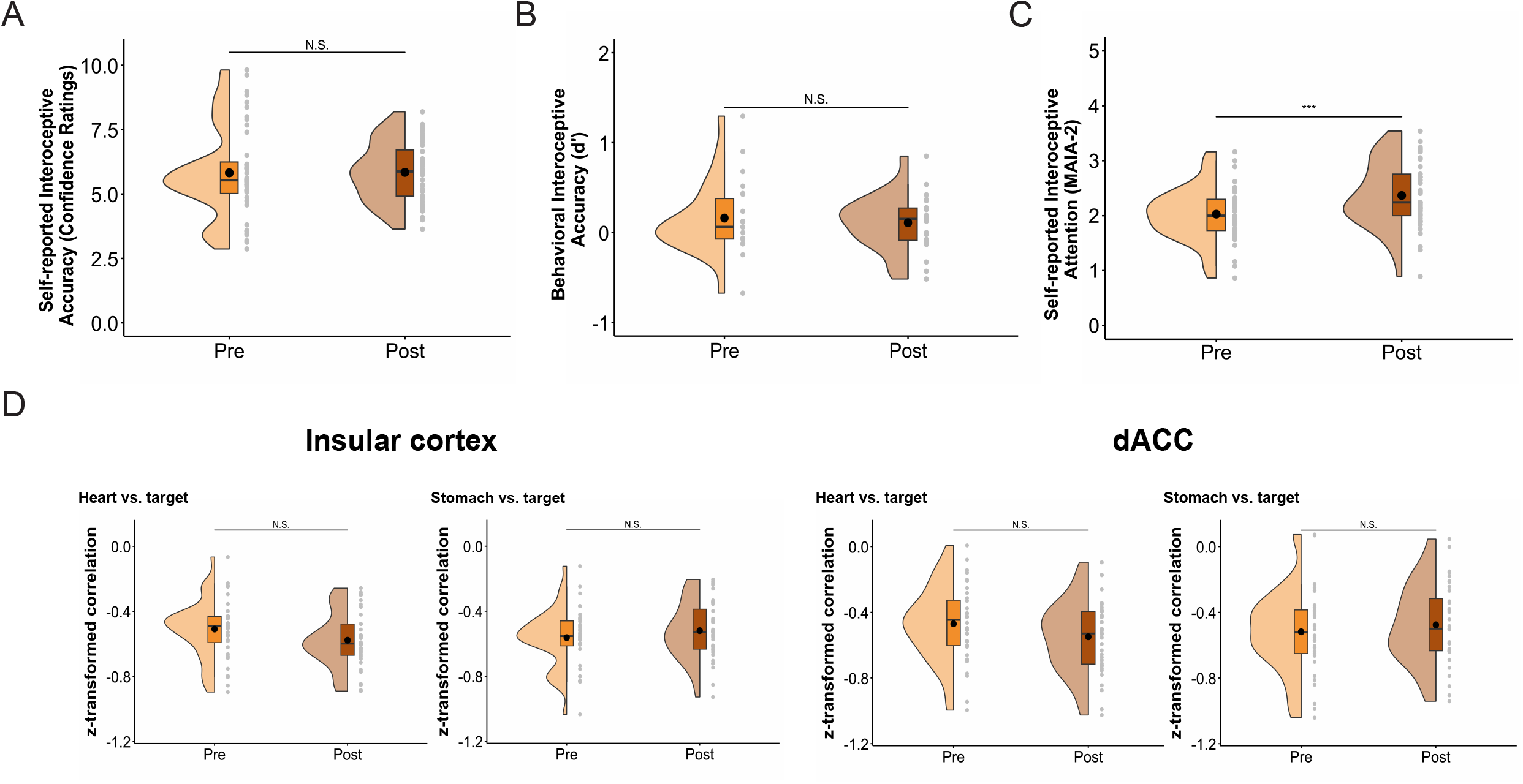
Effects of four-week Dialectical Behavior Therapy (DBT). Four weeks of DBT had no significant effect on self-reported (confidence ratings, **A**) or behavioral (d’, **B**) interoceptive accuracy. However, DBT was associated with a significant increase in self-reported interoceptive attention (MAIA-2 scores, **C**) in BPD patients (n=37). Furthermore, DBT had no significant effect on the neural representation of interoceptive versus exteroceptive attention in the insular cortex and the dACC (**D**). In the boxplot, the line dividing the box and the black dot represent the median and mean of the data. The ends represent the upper/lower quartiles, and the extreme lines represent the highest and lowest values, excluding outliers. Abbreviations: BPD, borderline personality disorder; \*\*\**p*<0.001; N.S., not significant. A Bonferroni-Holm correction was applied to adjust for multiple comparisons.

#### Interoceptive attention

Self-reported interoceptive attention improved after treatment (pre: 2.03 ± 0.50; post: 2.37 ± 0.61; *F*_(1,36)_ =22.87, *p*<0.0001, η _G_^2^=0.09; **Fig. 5C**, interaction effect time × group *F*_(1,66)_ =9.36, *p*=0.003, η _G_^2^=0.02). A comparison of pre- and post-fMRI data revealed no treatment-related changes within BPD patients and compared to HC for any contrast (all *p*_FWE_-values>0.05). Likewise, the multivariate approach showed no significant changes in neural representations within BPD patients after treatment (all *p*_(cor)_**-**values>0.05; **Fig. 5D**). In line with the neural results, there was no significant treatment-related effect on intensity ratings during interoceptive attention within BPD patients and compared to changes in HC (all *p*-values>0.05).

#### Moderation effects

Moderation analyses revealed a significant moderation effect of time × symptom change on both self-reported indicators of interoceptive attention (MAIA-2: *F*_(1,35)_ =8.15, *p*=0.007, η_p_^2^=0.19; intensity ratings: *F*_(1,34)_ =9.15, *p*=0.005, η_p_^2^=0.21). Johnson-Neyman plots indicated that both significantly increased after treatment (compared to pre-treatment) in patients who showed a symptom reduction of ≥0.11 or ≥1.17, respectively. There were no other significant moderation effects of changes in symptom severity (BSL-23 scores) or treatment response (responders vs non-responder). Likewise, correlations with changes in interpersonal problems (IIP scores) revealed no significant associations.

## Discussion

The study investigated interoception in BPD using a comprehensive, multifaceted approach as recommended by recent topical reviews (5–7). As hypothesized, BPD patients showed lower self-reported interoceptive attention and altered neural representation of cardiac interoceptive attention in the insular cortex and the dACC. Contrary to our hypotheses self-reported interoceptive accuracy and metacognitive awareness were not significantly altered in our BPD sample compared to HC, while behavioral interoceptive accuracy declined as BPD symptom severity increased. Importantly, only self-reported interoceptive attention improved after four weeks of a residential DBT program. These findings reveal a disrupted interoceptive profile in BPD, characterized by impairments in distinct interoceptive facets that respond differently to BPD treatment.

Our observation of lower self-reported interoceptive attention is consistent with previous studies reporting that BPD patients show reduced body ownership (38) and awareness (18). Importantly, reduced self-reported interoceptive attention was associated with interpersonal problems, a cardinal symptom of BPD (39). This underscores the clinical relevance of interoceptive dysfunction in BPD and supports the role of interoceptive attention for psychosocial deficits observed in BPD as proposed by an integrated biobehavioral model (10). Impaired interoceptive attention in BPD was further supported by objective measures as we observed altered neural representations of interoceptive attention to the heart in the insular cortex and the dACC, with BPD patients showing reduced differentiation between interoceptive and exteroceptive attention. This is in line with a pioneering study reporting that reduced cardiac interoceptive attention in BPD is associated with structural alterations in the insular cortex and dACC (13). Thus, these brain regions may be central to interoceptive dysfunction in BPD by disrupting the integration of interoceptive signals (13), particularly in relation to emotions (11, 40). This disruption may contribute to the previously hypothesized external attention bias (11, 14, 18), leading to emotional dysfunction and self-harming behavior as a maladaptive compensation (41). In the context of modern computational models of interoceptive psychopathology (42, 43), our findings indicate that altered interoception in BPD may result from prediction errors in the insular cortex, i.e., a mismatch between actual sensory input and expected body signals. These errors may arise from an overweighting of prior beliefs and experiences, such as a negative body image and an external attention bias, which in turn may lead to maladaptive response selection, including self-harm or intense emotional reactions. However, future studies are needed to evaluate this mechanistic account.

Interestingly, interoceptive accuracy seemed unaffected by this external attention bias, except in more severe BPD cases. A divergence between interoceptive attention and accuracy has been identified as clinically relevant in other conditions such as autism spectrum disorder (44) and may offer a starting point for targeted interventions. For instance, interoceptive training methods like ADIE (Aligning Dimensions of Interoceptive Experience) have been shown to enhance cardiac interoceptive accuracy, unlike mindfulness (45) or meditation practices (46), and can subsequently improve both interoceptive attention and anxiety (47). Within the framework of computational models (42, 43), interoceptive accuracy training may help to counterbalance an overweighting of negative priors and expectations by enhancing the processing of incoming sensory signals (48). This is particularly relevant for patients with more severe BPD, who may exhibit disruptions in interoceptive accuracy. Improvements in cardiac interoceptive accuracy can potentially result from an improved attentional switch from external to internal cues (49), a mechanism critical for a healthy state (6). Existing trainings on interoceptive accuracy could be adapted to train interoceptive attention by gradually adding distractors to match real-world conditions, as well as exteroceptive attention conditions, to train the attentional switch. In line with recent recommendations for interoceptive accuracy training (49), this approach could combine interoceptive attention training with non-interoceptive training (e.g., general attention training) to clarify whether improvements are interoception-specific or driven by a domain-general attention enhancement. Integrating such targeted training alongside interoceptive exposure and psychoeducation (49) into existing body-oriented therapy modules within DBT could be beneficial. Furthermore, smartphone-based applications could support and reinforce interoceptive attention skills between therapy sessions or during weekend stress trials, promoting the transfer of these skills to daily life.

Our study contributes to recent work in the field of interoception (5–7) by emphasizing the importance of simultaneous assessment of multiple facets of interoception. It is important to note, however, that our findings are interpreted within the framework proposed by Murphy and colleagues (5), whereas other models of interoception encompass additional facets (6, 7). Only by applying a comprehensive, multifaceted approach it is possible to identify how different interoceptive facets are uniquely impaired and selectively respond to treatment. For example, the significant treatment effect on self-reported interoceptive attention, despite the absence of changes in other interoceptive facets, may reflect the strong focus of DBT on mindfulness with emphasis on awareness (i.e., non-judgmental awareness of internal experiences) (8, 50) in combination with cognitive reappraisal (51). Our findings highlight a potential therapeutic benefit of additional interoceptive attention training for all BPD patients, and accuracy training for more severe cases. However, it is crucial to note that interoceptive accuracy training can also have adverse effects (48, 49, 52), further underscoring the importance of isolating selective interoception impairments to tailor treatment recommendations effectively. As accuracy (53) and mindfulness training (20–23) may also impact interoception-related neural networks, further fMRI longitudinal studies are needed. Lastly, our finding of reduced differentiation of interoceptive and exteroceptive attention to the heart but not the stomach additionally underscores the importance of examining interoception across different bodily axes (54).

The present study has some limitations. We recruited a naturalistic cohort of BPD patients who presented with comorbidities and were under psychotropic medication. A previous study has shown that while neural alterations during interoceptive attention did not differ between medicated and unmedicated MDD patients, intensity ratings were significantly higher in medicated patients (55). To address this, we controlled for the potential effects of medication by including psychotropic medication use as a covariate in our analyses. Additionally, the heartbeat discrimination task proved very challenging even for the HC group. Detecting subtle differences between HC and less severe BPD patients may require more sensitive tasks. Furthermore, our study lacks a waiting-list patient group, and interoception-related treatment changes may have become evident after the completion of all three DBT modules.

To conclude, this study demonstrates that BPD is characterized by disturbances in specific interoceptive facets, which differently respond to treatment. As such, it highlights the importance of comprehensive, multifaceted assessments of interoception in clinical conditions and the therapeutic potential of interoceptive attention training for all BPD patients, with accuracy training for more severe cases.

## Acknowledgment

D.S. was supported by a Research pool University of Oldenburg Medical Scientists grant (FP 2020-047).

The Center for Magnetic Resonance Research (CMRR) sequence was kindly provided by the University of Minnesota Center for Magnetic Resonance Research. Preprocessing of fMRI data was performed on the HPC Cluster CARL funded by the DFG under INST 184/157-1 FUGG. We thank the patients and the DBT team, particularly Gudrun Hemje-Oltmanns and Lara Preis. We also express our gratitude to Hannah Allmandinger, Paulina Piwkowski, Marlene Charlotte Holzhausen, Nick Michalek, Paul Leonard Grupe, and Anja Sablotny for their assistance with data collection.

## Author Contributions

J.V., D.S., D.P., and C.M. designed the experiments; J.V. conducted the experiments; J.V., D.P., and D.S. analyzed the data. J.V., D.S., D.P., C.M., M.R., R.H., A.S. and R.B.-B. wrote the manuscript. All authors read and approved the manuscript in its current version.

## Data availability

The data of this study are not publicly available due to privacy reasons. The data that support the findings of this study are available from the corresponding author upon reasonable request and with a data sharing agreement in place.

