## Supplementary Information for "Selective interoception impairments and treatment effects in Borderline Personality Disorder"

Voelter et al.	Supplement

***Borderline Personality Disorder***

**Supplementary methods**

*Power-analysis*

We used G*Power 3 (1) to conduct an a-priori power analysis for the study based on the effect size reported in a Cochrane Review on psychological therapies for patients with borderline personality disorder (BPD) (2). The authors observed an overall medium effect size of *d* = -0.6 for dialectical behavior therapy (DBT) treatment effects on borderline symptom severity. A different study (3) found the same effect size (*d* = -0.6) for positive effects of DBT treatment on interpersonal problems in BPD. Because the study focuses on the effects of one DBT module (four weeks treatment), we assumed a reduced effect size of *d* = 0.5. To replicate the abovementioned DBT treatment effect on borderline symptom severity and interpersonal problems in BPD (with *α* = 0.05 and power = 0.9, one-tailed paired-sample t-test) at least 36 patients have to be tested. Assuming a possible drop-out rate of 20-25% due to the longitudinal study design, we planned to test at least 50 patients.

A previous study (4) observed large effect sizes for differences in social functioning between BPD patients and HC. Assuming a large effect (*d* = 0.8) and considering an allocation ratio of 2:1, based on the assumption of a 50% treatment response rate in the patient group, (with *α* = 0.05 and power = 0.9, two-tailed two-sample t-test), at least 25 HC have to be tested. To account for possible drop-outs of 10-15%, we planned to test at least 30 HC.

*Residential DBT program*

The residential DBT at the Karl-Jaspers-Klinik includes the following treatment components:

- Individual therapy (2 hours per week)
- Group skills training (2 hours per week)
- Group psychoeducation (1 hour per week)
- Peer group meetings (2 hours per week)
- Mindfulness group (1 hour per week)
- Individual body-oriented therapy (1.5 hours per week)
- Therapist team consultation meetings (2 hours per week)

The psychoeducation sessions offer insight into Marsha Linehan’s biosocial theory of BPD, complemented by the latest theoretical developments and research findings on BPD. The mindfulness group builds upon the standard DBT skills training mindfulness module, with a stronger focus on sitting practice compared to traditional outpatient DBT. Meanwhile, the body-oriented therapy consists of education on psychomotor interactions and exercises aimed at enhancing body awareness and perception (3).

The residential DBT program at the Karl-Jaspers-Klinik is led by trained psychologists and is supported by medical doctors and nurses. It has been officially certified by the DBT Dachverband (DBT e.V.) since 2011. The staff of the DBT ward follows the German interactive skills manual for borderline patients by Martin Bohus and Martina Wolf-Arehult (5). This manual is based on the DBT skills training by Marsha Linehan (6). The content has been revised and expanded in cooperation with Marsha Linehan. It includes essential background information and instructions for conducting group skills training, as well as concrete worksheets and informational materials. In addition to the treatment sessions mentioned above, BPD patients begin their day with a morning round, where they discuss their plans and goals for the day. They end each day by reviewing their diary card with the nurses, which tracks daily events and tension curves throughout the day. Furthermore, at the Karl-Jaspers-Klinik, patients participate in additional therapeutic activities which are an integrated part of the DBT and vary depending on individual needs:

- Ergotherapy
- Dance and theater therapy
- Woodworking workshops

Patients engage in these treatments approximately about six times per DBT module. Additionally, patients meet once a week with the chief senior physician to discuss medication adjustments and with their supervising psychiatric nurse to talk about any problems during daily living.

DBT at the Karl-Jaspers-Klinik comprises three modules, each followed by a break of varying duration. Patients were recruited from all three DBT modules (1^st^ module n=23, 2^nd^ module n=10. 3^rd^ module n=4).

*Participants*

General exclusion criteria were age under 18 or over 65 years, MRI contraindication, scars on a predefined area of 20 cm of their shins, acute suicidality, any lifetime psychotic disorders, current substance dependence, a history of traumatic brain injuries, or other neurological illnesses. The presence of a mental disorder or current or past psychiatric inpatient treatment resulted in exclusion from study participation in the HC group.

Axis I disorders were assessed by the Structured Clinical Interview for DSM-5 Disorders: clinician version (SCID-5-CV) (7). HC additionally underwent the Zanarini Rating Scale for Borderline Personality Disorders (ZAN-BPD) (8) and completed the Assessment of DSM-IV Personality Disorders (ADP-IV) (9) to confirm the absence of any current mental disorders. The average time between the last study appointment and admission to DBT was 4.15 ± 5.10 weeks. Accordingly, the HC group was assessed with a waiting period of five to nine week between pre- and post-measurement.

*Psychological and clinical assessments*

The Beck Depression Inventory-II (BDI-II) (10) and the Liebowitz Social Anxiety Scale (LSAS) (11) were used to measure depressive symptoms and social anxiety, respectively. Inner tension was assessed on a visual analog scale (VAS) from 0 (not at all agitated) to 100 (very agitated) approximately half an hour before participants entered the MRI scanner. Furthermore, hunger and the urge to urinate were evaluated verbally immediately before the visceral interoceptive awareness (VIA) task on a scale from 0 (no hunger/urge to urinate) to 100 (very strong) while the participant was inside the MRI. Except for the Childhood Trauma Questionnaire (CTQ), all participants completed the questionnaires before and after four weeks of residential DBT/waiting interval. Depressive symptoms and social anxiety were only analyzed at baseline to characterize the sample.

*Heartbeat discrimination task*

To minimize noise disturbances, the experimenter left the room and closed the door and windows. Participants were instructed not to manipulate their heart rate (e.g., by holding their breath) or measure their pulse manually. The experiment required the detection of at least three consecutive, artifact-free heartbeats to initiate a tone sequence. The task comprised 80 trials, with 40 trials featuring asynchronous tones (randomly delayed by 550 ± 75 ms) and 40 trials featuring synchronous tones (randomly delayed by 250 ± 75 ms). Task duration varied based on participants' reaction time but generally ranged from 20 to 25 minutes. The heartbeat discrimination task was applied before and after one module (i.e., four weeks) of the residential DBT program.

*Visceral interoceptive awareness (VIA) task*

Each of the three conditions (heart, stomach, target) consisted of 12 trials, resulting in a total of 36 trials and ~14 minutes of measurement time. The task was divided into two runs of 18 trials each, separated by a 30-second break. Each rating lasted five seconds. To optimize trial arrangements (order of conditions and duration of interstimulus intervals (ISIs), which were randomized between 2.5 and 12.5 seconds), the easy-optimize-x implementation for MATLAB (The MathWorks, Natick, MA) by Bob Spunt (https://www.bobspunt.com/easy-optimize-x/) was used. Optimal designs were generated and randomly assigned to participants, with variations between pre- and post-measurement. MRI data was acquired using a 3 Tesla Siemens MAGNETOM Prisma Scanner (Siemens AG, Erlangen, Germany) with a Siemens 64-channel head coil. For fMRI data, a T2*-weighted echoplanar (EPI) multiband sequence with a multiband acceleration factor of four was used to measure neural responses to interoceptive attention (Repetition time (TR) = 850 ms, Echo Time (TE) = 30 ms, matrix size: 76 x 76, voxel size: 2.5 x 2.5 x 2.5 mm³, slice thickness = 2.5 mm, distance factor = 0 %, field of view (FoV) = 192 x 192 mm², flip angle 62°, 48 slices). To control for inhomogeneities of the magnetic field, a fieldmap was obtained prior to the VIA task and was included during preprocessing of the fMRI data (TR = 533 ms, TE (1) = 5.19, TE (2) = 7.38, matrix size: 64 x 64, voxel size: 3 x 3 x 3 mm³, slice thickness = 3.0 mm, distance factor = 33 %, FoV = 192 x 192 mm², flip angle 60°, 35 slices). High-resolution T1-weighted structural images were collected at the same scanner (TR = 2000 ms, TE = 2.07 ms, matrix size: 320 x 320, voxel size: 0.8 x 0.8 x 0.8 mm³, slice thickness = 0.75 mm, FoV = 240 x 240 mm², flip angle = 9°, 224 slices) and were included during the analysis of fMRI data. The fMRI VIA task was performed before and after one module (i.e., four weeks) of the residential DBT program.

*Behavioral data analysis*

The statistical analyses were conducted using R (12) and MATLAB release R2021a (The MathWorks, Natick, MA).

To assess possible group effects on confidence ratings of the heartbeat discrimination task, mixed-design Analyses of Covariance (ANCOVAs) including the between-subject factor group (HC, BPD), the within-subject factor trial type (incorrect, correct; e.g. the synchronous tones were recognized as synchronous in correct trials) and covariates were computed. To assess treatment-related effects, the within-subject factor time (pre, post) and the interaction of group and time were additionally included.

A mediation analysis with childhood maltreatment (CTQ scores) as predictor, self-reported interoceptive attention (MAIA-2 scores) as mediator, and interpersonal problems (IIP scores) as an outcome was conducted to examine the relationship between adverse childhood environments, resulting psychosocial deficits, and interoception as an underlying mechanism.

To measure the effect sizes, we employed Cohen's d, partial eta squared (η_p_^2^), and generalized η² (η_G_²) (13, 14). To determine the magnitude of the effects, the following commonly used benchmarks can be applied (13): for Cohen's d, a small effect is indicated by *d* = 0.2, a medium effect by *d* = 0.5, and a large effect by *d* = 0.8. Similarly, for both η_p_^2^ and η_G_², the benchmarks are as follows: a small effect corresponds to η_p_^2^ = 0.01, a medium effect to η_p_² = 0.06, and a large effect to η_p_^2^= 0.14.

*fMRI data analysis*

fMRI data were preprocessed using the standardized pipeline *fMRIPrep* 20.2.1 (15), which is based on *Nipype* 1.5.1 (16, 17). The first three volumes of each functional time series were discarded to allow for T1 equilibration. The pipeline included correction for magnetic field inhomogeneities and removal of movement components using Independent Component Analysis - Automatic Removal of Motion Artifacts (ICA-AROMA). Additionally, we checked whether the standardized derivative of the root mean squared variance over voxels (std. DVARS) of our participants exceeded a threshold of 1.5. This was the case for zero participants. Physiological noise was corrected using the Component Based Noise Correction Method (CompCor). The fMRI analysis was conducted using SPM12 (https://www.fil.ion.ucl.ac.uk/spm/), implemented in MATLAB release R2021a (The MathWorks, Natick, MA). The analysis involved a two-level approach, analyzing the data on single-subject and group level. General Linear Models (GLMs) were used, and GLM parameters were estimated using the restricted maximum likelihood method. The FAST model was applied to correct for autocorrelations. Hemodynamic responses to all three conditions (heart, stomach, target) were modeled as boxcar functions with condition duration as length. The duration of ratings, the break between runs, three cerebrospinal fluid (cfs) and two white matter (wm) CompCor components explaining the most variance in the data and cosine regressors as a high-pass filter were included as nuisance regressors.

The fMRI analysis in SPM12 involved a two-level approach based on the General Linear Model. Treatment effects were investigated by calculating differences between pre- and post-measurement data in BPD patients and HC on the first level. Second-level statistical inference included two-sample and one-sample t-tests. To define the regions of interest (ROIs), we used the Human Brainnetome Atlas (18) and merged them into a single ROI mask using the BRAinNetome Toolkit (BRANT) (19) to adjust for multiple ROIs. The individual brain regions consist of the following subregions: vmPFC (41 OrG_L_6_1, 42 OrG_R_6_1 45, OrG_L_6_3, 46 OrG_R_6_3 OrG_L_6_4, 48 OrG_R_6_4, 49 OrG_L_6_5, 50 OrG_R_6_5), amygdala (211 Amyg_L_2_1, 212 Amyg_R_2_1, 213 Amyg_L_2_2, 214 Amyg_R_2_2), dACC (179 CG_L_7_3, 180 CG_R_7_3, 183 CG_L_7_5, 184 CG_R_7_5) and the insular cortex (163 INS_L_6_1, 164 INS_R_6_1, 165 INS_L_6_2, 166 INS_R_6_2, 167 INS_L_6_3, 168 INS_R_6_3, 169 INS_L_6_4, 170 INS_R_6_4, 171 INS_L_6_5, 172 INS_R_6_5, 173 INS_L_6_6, 174 INS_R_6_6).

To validate that the fMRI VIA task elicited activation in an interoception-related brain network, we analyzed data of HC at baseline using the ROI mask comprising the insular cortex, dACC, amygdala, and vmPFC with a one-sample t-test on the contrast interoception>exteroception.

*fMRI representational similarity analyses (RSAs)*

All ROI-based RSAs were computed using The Decoding Toolbox (TDT) (20), and bootstrapped t-tests were calculated with 10,000 replicates.

*Missing values*

Five BPD patients have missing data on years of education, and two BPD patients did not complete the MAIA-2 for the pre-measurement. For one BPD patient, we imputed the covariates hunger and urge to urinate using the mean of the group for the fMRI VIA task at baseline. Due to technical problems, baseline assessments of the heartbeat discrimination task were available only for 49 BPD patients and 26 HC, resulting in longitudinal data from 26 BPD patients and 26 HC.

Consistent with our preregistration (<https://osf.io/htf2n>), the collected variables and analyses remained unchanged; however, variable labels were adjusted to match the terminology of the more appropriate model by Murphy et al (21).

As part of a larger study, additional variables were assessed before, after, and during residential treatment (reported elsewhere).

**Supplementary results**

*Questionnaire data*

To determine whether the significant correlation between self-reported interoceptive attention (MAIA-2 scores) and interpersonal problems (IIP scores) was driven by an outlier in the patient group, we recalculated the correlation excluding that subject. The correlation remained significant within BPD patients (*r*_(50)_=-0.42, *p*_(cor)_=0.004) and across both groups (*r*_(50)_=-0.62, *p*_(cor)_<0.0001).

*Heartbeat discrimination task*

Before the analysis, all trials from the heartbeat discrimination task were plotted and screened for noise. Noisy trials were identified and excluded from further analysis. If the number of noisy trials exceeded 20% of all trials (i.e., more than 16 trials), the entire subject was excluded from further analysis. This procedure resulted in the exclusion of 9 BPD patients and 0 HC from the baseline analysis and an additional 4 BPD patients and 3 HC from the analysis of DBT effects.

The mixed-design ANCOVA with group as between-subject factor (HC, BPD) and trial type (correct, incorrect) as within-subject factor revealed no significant group effect nor an interaction effect with group on confidence ratings of the heartbeat discrimination task (all *p*-values > 0.05). Additionally, there were no treatment-related effects on confidence ratings within BPD patients and compared to HC (all *p*-values > 0.05).

The d’-values in the HC group clustered around zero, suggesting that the task was very challenging. However, we tested whether d’-values of the HC group were significantly different from zero using a one-sample t-test. Mean d’-values of HC (0.29±0.41) were significantly higher than zero (*t_(25)_*= 3.68, *p*=0.001), indicating above-chance performance. Similarly, BPD patients also showed mean d’values (0.20±0.47) that were significantly different from zero (*t_(39)_*= 2.77, *p*=0.009).

Some BPD patients exhibited negative d′ values, indicating performance below chance level. To account for the potential influence of these values on the moderation effect of symptom severity (BSL-23 scores) on behavioral interoceptive accuracy, we set all negative d′ values to zero and repeated the moderation analysis. The results showed that the moderation effect of BSL-23 scores × group on behavioral interoceptive accuracy remained significant (*F*_(1,58)_=10.50, *p*=0.002*,* η_p_^2^=0.15).

*Mediation analysis*

A mediation analysis using childhood maltreatment (CTQ scores) as a predictor, self-reported interoceptive attention as a mediator (MAIA-2 scores), and interpersonal problems (IIP scores) as the outcome measure further revealed a significant direct effect of childhood maltreatment on interpersonal problems (*B*=0.01, *p* < 0.0001) but no significant mediation effect (*B*=0.001, *p*=0.43) of self-reported interoceptive attention on this relation.

*Intensity ratings of the VIA task*

The mixed-design ANCOVA with group as between-subject factor (HC, BPD) and condition (stomach, heart, target) as within-subject factor revealed no significant group effect nor an interaction effect with group (all *p*-values > 0.05) but a significant main effect of condition (*F*_(2,156)_=7.64*, p*<0.01*,* η_G_^2^*=*0.06) on the intensity ratings of the VIA task. As expected, the exteroceptive condition target (HC: 6.04 ± 1.74, BPD: 6.19 ± 1.68) was rated as more intense than the interoceptive conditions stomach (HC: 5.65 ± 1.50, BPD: 4.47 ± 2.19) and heart (HC: 5.29 ± 1.71, BPD: 5.05 ± 1.79) in both groups (stomach vs. target: *t*_(79)_=-4.38, *p*_(cor)_=0.0001; heart vs. target: *t*_(79)_=-3.36, *p*_(cor)_=0.002)*.*

*Exploratory whole brain level analysis*

An exploratory whole brain level analysis revealed group differences in the left supplementary motor area for interoceptive attention to the stomach relative to exteroceptive attention (MNI: -10, 4, 56; *t*_(72)_=4.84*; p*_FWE_=0.011) prior to treatment. Further, activity of the right inferior frontal gyrus, triangular part changed significantly after treatment within BPD patients for attention to the heart compared to exteroception (MNI: 36, 28, 28; *t_(_*_26)_=7.71*; p*_FWE_=0.046). There were multiple changes in neural activity of different brain regions in BPD patients compared to HC after treatment (see **Supplementary Tab. 4**). We further detected a significant treatment response × time interaction effect in the right precuneus (MNI: 6, -46, 58; *t*_(25)_=5.10*; p*_FWE_=0.004) for interoceptive attention versus exteroceptive attention after treatment within BPD patients. Significance was assessed at cluster level with *p* < 0.05, FWE corrected and brain regions were identified with the Automated Anatomical Labeling (AAL) Atlas (22, 23).

*RSA bootstrapping*

As exact bootstrapped *p*-values vary slightly across iterations, we report multiple comparison–corrected bootstrapped *p*-values (*p*_(corBoot)_) from a representative iteration. These values reflect comparisons of neural representations during cardiac interoceptive attention versus exteroceptive attention in BPD patients compared to HC. Specifically, BPD patients exhibited greater similarity in neural activity patterns between interoceptive attention to the heart and the exteroceptive condition, characterized by significantly reduced negative correlations compared to HC in both regions (insular cortex: BPD: *r*_(48)_=-0.51, HC: *r*_(29)_=-0.61, *t*_(79)_=3.18, *p*_(corBoot)_=0.007, *d*=0.69; dACC: BPD: *r*_(48)_=-0.49, HC: *r*_(29)_=-0.61, *t*_(79)_=2.46, *p*_(corBoot)_=0.046, *d*=0.56).

*RSA results control analysis*

As preregistered, bootstrapped t-tests were used to examine differences in neural representations of interoceptive attention between BPD patients and HC. To assess whether the observed group differences in neural activation within the insular cortex and the dACC remained significant after controlling for potential confounding variables (age, sex, BMI, psychotropic medication use, hunger, the urge to urinate, and prescan inner tension), we conducted additional ANCOVAs. These models included group (HC, BPD) and the covariates as predictors, with the z-transformed correlation coefficients of the RSAs as the dependent variable. Results from both ANCOVAs indicated that the group differences remained statistically significant after adjusting for covariates (main effect of group insular cortex: *F*_(1,72)_=7.95, *p*=0.006, η_p_^2^*=*0.10; dACC: *F_(_*_1,72)_=6.25, *p*=0.02, η_p_^2^*=*0.08).

*Additional exploratory analyses*

The z-transformed correlation coefficients of the RSAs were not significantly associated with the intensity ratings of the VIA task, averaged across interoceptive conditions (all *p*-values > 0.05).

There was a trend towards a significant moderation effect of group × BSL-23 scores on confidence ratings, averaged across correct and incorrect trials of the heartbeat discrimination task (*F_(_*_1,58)_=3.71, *p*=0.059).

**Supplementary Figures**

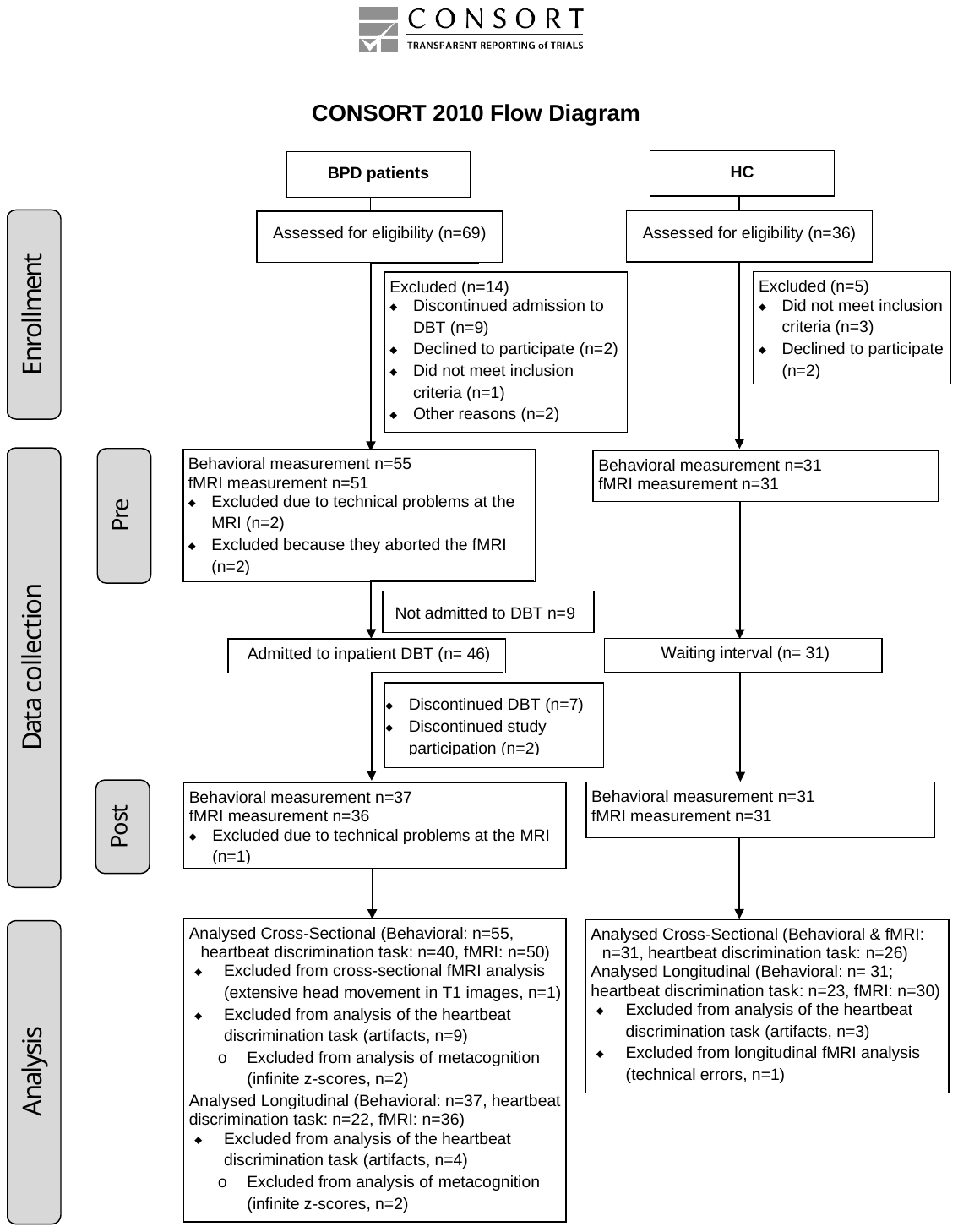

**Supplementary Figure S1: CONSORT flow diagram of the entire study process.**

**Supplementary Tables**

**Supplementary Table S1: Sample description for patients with BPD and HC.** The higher number of women than men reflects the distribution of BPD patients treated in clinical settings (24). All study participants are Caucasian.

HC (n=31)

BPD patients (n=55)

| Characteristics | M ± SD  or n | Min/Max | M ± SD  or n | Min/Max | X^2^ / t-test,  p |
| --- | --- | --- | --- | --- | --- |
| Demographic data |  |  |  |  |  |
| Age, years | 28.3 ± 10.2 | 18/57 | 28.0 ± 10.9 | 18/60 | 0.9 |
| Sex, male/female | 7/48 |  | 4/27 |  | 1 |
| Gender, male/female/diverse | 6/47/2 |  | 4/27/0 |  |  |
| BMI | 29.6 ± 7.1 | 16.2/45 | 24.2 ± 4.5 | 18.8/36.6 | <0.0001 |
| Education, years | 13.6 ± 2.8* | 9/24 | 17.3 ± 3.0 | 13/24 | <0.0001 |
| Psychological data |  |  |  |  |  |
| Depression (BDI-II) | 37.2 ± 12.5 | 6/57 | 3.6 ± 3.7 | 0/15 | <0.0001 |
| Social anxiety (LSAS) | 76.3 ± 30.6 | 1/124 | 24.5 ± 11.5 | 4/44 | <0.0001 |

### *Notes. BDI-II, Beck Depression Inventory-II; BMI, Body Mass Index; BPD, borderline personality disorder; HC, healthy controls; LSAS, Liebowitz Social Anxiety Scale; M, Mean; n, number of available data points; SD, Standard Deviation; * data not available for 5 BPD patients*

**Supplementary Table S2: Axis I disorders and psychotropic medication of BPD patients and HC.**

HC

BPD patients

| Characteristics | n | n |
| --- | --- | --- |
| Axis I disorders (DSM-V criteria) |  |  |
| Any current comorbid diagnosis, yes/no | 48/7 | 0/31 |
| Any lifetime comorbid diagnosis, yes/no | 53/2 | 0/31 |
| Psychotropic drugs |  |  |
| Antidepressants | 35 | 0 |
| Antipsychotics | 22 | 0 |
| Psychostimulants | 6 | 0 |
| Anticonvulsants | 2 | 0 |

### *Notes. BPD, borderline personality disorder; HC, healthy controls; n, number of available data points*

**Supplementary Table S3: Longitudinal sample description for patients with BPD and HC.**

HC (n=31)

BPD patients (n=37)

| Characteristics | M ± SD  or n | Min/Max | M ± SD  or n | Min/Max | X^2^ / t-test,  p |
| --- | --- | --- | --- | --- | --- |
| Demographic data |  |  |  |  |  |
| Age, years | 28.1 ± 10.7 | 19/57 | 28.0 ± 10.9 | 18/60 | 1 |
| Sex, male/female | 5/32 |  | 4/27 |  | 1 |
| Gender, male/female/diverse | 4/32/1 |  | 4/27/0 |  |  |
| BMI | 29.7 ± 6.6 | 16.2/45 | 24.2 ± 4.5 | 18.8/36.6 | <0.001 |
| Education, years | 14.2 ± 2.7* | 10/24 | 17.3 ± 3.0 | 13/24 | <0.001 |
| Psychological data |  |  |  |  |  |
| Depression (BDI-II) | 40.0 ± 9.4 | 6/57 | 3.6 ± 3.7 | 0/15 | <0.0001 |
| Social anxiety (LSAS) | 79.9 ± 29.6 | 1/124 | 24.5 ± 11.5 | 4/44 | <0.0001 |

### *Notes. BMI, Body Mass Index; BPD, borderline personality disorder; HC, healthy controls; M, Mean; n, number of available data points; SD, Standard Deviation; * data not available for 2 BPD patients*

**Supplementary Table S4: Whole brain results for treatment-related changes of BPD patients compared to HC.**

| 1^st^ level contrast | 2^nd^ level contrast | MNI  coordinates | t-score | p_FWE_ | Brain region |
| --- | --- | --- | --- | --- | --- |
| Pre _interoception > target_ >  Post _interoception > target_ | BPD > HC | -38 8 52 | 5.24 | 0.011 | Left middle frontal gyrus |
|  |  | 16 -72 16 | 4.61 | >0.0001 | Right calcarine fissure and surrounding cortex |
|  |  | 0 -80 -8 | 4.43 | >0.0001 | Left calcarine fissure and surrounding cortex |
|  |  | 22 -92 24 | 4.10 | 0.026 | Right superior occipital gyrus |
|  |  | -10 -74 26 | 4.01 | 0.034 | Left cuneus |
| Pre _stomach > target_ >  Post _stomach > target_ | BPD > HC | 18 -68 8 | 4.84 | 0.001 | Right calcarine fissure and surrounding cortex |
|  |  | -16 76 -12 | 4.17 | 0.002 | Left lingual gyrus |
| Pre _heart > target_ >  Post _heart > target_ | BPD > HC | -40 8 52 | 5.14 | 0.004 | Left middle frontal gyrus |
|  |  | 8 -28 4 | 5.11 | 0.006 | Right thalamus |
|  |  | -6 -16 6 | 5.10 | 0.009 | Left thalamus |
|  |  | 0 -80 -8 | 4.59 | >0.0001 | Left lingual gyrus |
|  |  | -24 -78 -14 | 4.28 | 0.003 | Left lingual gyrus |

*Notes. BPD, borderline personality disorder; HC, healthy controls; MNI, Montreal Neurological Institute; FWE, family-wise error corrected*
